# Relative assessment of cloth mask protection against ballistic droplets: a frugal approach

**DOI:** 10.1101/2022.03.10.22272219

**Authors:** V. Márquez-Alvarez, J. Amigó-Vera, A. Rivera, A. J. Batista-Leyva, E. Altshuler

## Abstract

During the COVID-19 pandemic, the relevance of evaluating the effectiveness of face masks –especially those made at home using a variety of materials– has become obvious. However, quantifying mask protection often requires sophisticated equipment. Using a frugal stain technique, here we quantify the “ballistic” droplets reaching a receptor from a jet-emitting source which mimics a coughing, sneezing or talking human –in real life, such droplets may host active SARS-CoV-2 virus able to replicate in the nasopharynx. We demonstrate that materials often used in home-made face masks block most of the droplets. Mimicking situations eventually found in daily life, we also show quantitatively that less liquid carried by ballistic droplets reaches a receptor when a blocking material is deployed near the source than when located near the receptor, which supports the paradigm that your face mask does protect you, but protects others even better than you. Finally, the blocking behavior can be quantitatively explained by a simple mechanical model.

## INTRODUCTION

More than two years have passed since start of the COVID-19 outbreak. From the early beginnings, face masks have had a key role in cutting off the transmission together with other preventive measures such as quarantines, physical distancing and hand hygiene. Many studies assessed masks’ efficacy [1-23], which compelled almost every health agency, including WHO [24], CDC [25] and ECDC [26], to recommend the use of face masks in certain settings.

By the end of 2020, the first vaccines were available [27], which has received a great deal of attention. While more than half of world population has been fully vaccinated by the time of writing this paper [28], the number goes below 6% in the case of low-income countries, representing a total population of 665 million people [29]. In fact, it seems “unlikely” that people from low-income countries will be fully vaccinated by the end of 2022 [30].

Moreover, evidence suggests that new variants of the SARS-CoV-2 is substantially more transmissible than previous strains of the virus [31]. Also, vaccinated individuals infected with some of them could carry the same amount of viral load than an unvaccinated person, meaning that they can also transmit the virus [32]. These findings have eventually made CDC to update its guidance and recommend wearing a mask in public indoor places, in areas of substantial or high transmission risk, even if they are fully vaccinated [33].

So, in the present context, face masks still play a central role in the fight against the pandemic. This includes home-made cloth masks, which coexist with commercial ones (like surgical and N95 masks) in countries like Cuba.

Evidence suggests that SARS-CoV-2 is transmitted mainly through direct exposure to respiratory droplets carrying the virus [34]. This respiratory transmission route is usually split in two ways. The first –that we will call *ballistic transmission*– occurs by the emission of large droplets of fluid as infected individuals sneeze, cough, sing, or talk. Those droplets – which follow ballistic trajectories barely unaffected by gas flows– can get in touch with the mucous membranes (eyes, nose or mouth) of a susceptible person and infect her. Otherwise they fall to the ground within 1-2 meters in the horizontal direction. The second –*aerosol transmission*– is linked to fine droplets, which, thanks to Brownian motion, can be suspended in the air for hours and easily travel with air currents. In real life this separation is not perfectly sharp, but varies continuously from one transmission route to another [34,35].

The relevance of aerosol transmission has been a subject of heated debate during the pandemic. Some studies conclude that aerosol transmission is plausible [35-39], while others agree, but argue that it involves low risk [40, 41], since before the 2-meter (∼ 6-feet) length scale, large droplets carry more viral load than airborne particles. Beyond that distance, aerosol droplets dilute in the air, and are easily carried by air currents (except in poorly ventilated places where increasing viral load concentration could make infection possible). In fact, the strong dependence of COVID-19 infection risk with people proximity suggests that ballistic transmission is more relevant than aerosol transmission [34].

Beyond the discussion about the dominant transmission mechanisms, it is safe to say that the generalized use of face masks provides protection in two ways: by limiting the emission of droplets from an infected subject into the environment (source control), and by reducing the inhalation and deposition of droplets by the wearer (wearer protection).

The present study has two main objectives: (1) Introducing a frugal experimental setup that allows to compare quantitatively the protection from ballistic droplets by different porous cloths used in the fabrication of home-made masks and (2) Testing the system in situations involving close encounters where individuals are wearing or not face masks, or are using them incorrectly.

By means of controlled experimentation using affordable materials and equipment, we evaluate the blocking capacity of a few materials commonly used in home-made face masks, which are popular in countries like Cuba. In the process of studying realistic situations using our setup, we show quantitatively that a blocking material deployed very near a source of ballistic droplets protects from them a receptor located farther away better than the same material deployed very near the receptor.

## EXPERIMENTAL

We horizontally spray blue-colored water on a screen: a nozzle producing the spray plays the role of the mouth or nose of a sneezing, coughing, or talking individual (source), while the screen plays the role of the face of a second individual (receptor). Between the two, an obstacle is deployed, consisting in a flat piece of porous fabric. The blocking element plays the role of a facial cloth mask. Since the droplets travel inside a plastic bucket, external air currents do not perturb the experiment. The spray is shot by pulling down the nozzle using a u-shaped thin wire attached to two levers coupled to servomotors. They are controlled by an Arduino Uno platform allowing to manually trigger the nozzle by pushing a micro-switch, in order to achieve reproducibility and decrease spurious vibrations that would occur if the nozzle was directly pushed down by hand. Our model experiment matches reasonably well the temporal evolution of the particle front velocity associated to coughing humans [43, 44] (see Fig. S1 –Multimedia view– in supplementary material). Fig.1(a) shows a sketch of the experimental device: a cloth is deployed at a distance of 1.5 cm from the screen, while the distance from nozzle to screen is 19.0 cm, which will be called Configuration 1 (CONF1). We will call Configuration Free (CONFFREE) the case in where there are no blocking materials between the nozzle and the screen. Fig. 1(b) shows a photograph of the device.

**Figure 1.**
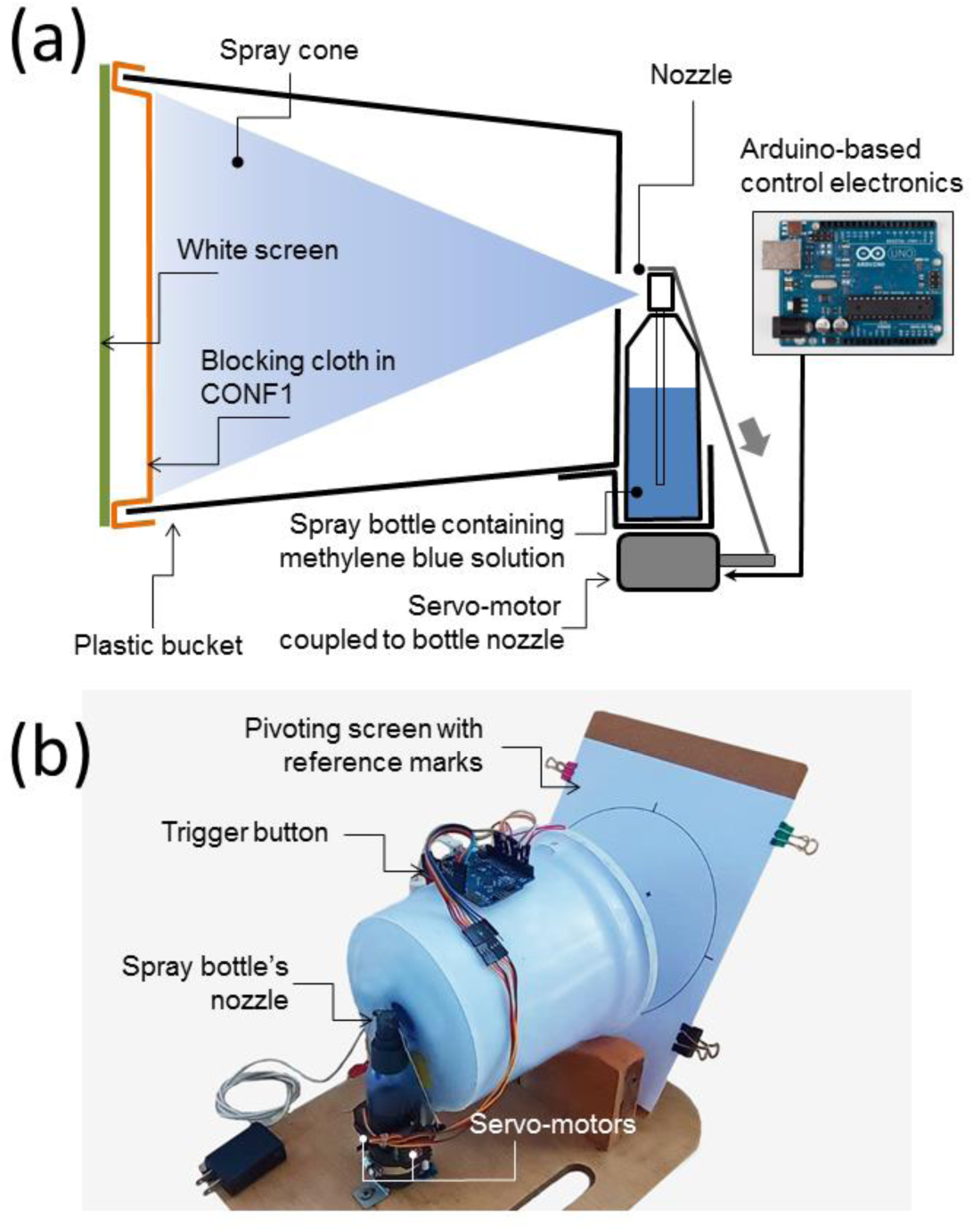
Experimental setup. (a) Sketch of the experimental setup, illustrating a cloth deployed in CONF1 (i.e., located 1.5 cm apart from the screen). (b) External photograph of the real device. The device is partially open so the screen can be seen, but not the blocking cloth.

Each experiment resulted in a pattern of blue stains on a white surface, which was digitally scanned and binarized, so clear and black areas correspond to places hit or not hit by droplets, respectively. If the screen is deployed at a distance of 4 cm from the nozzle with no blocking material in between, the resulting stain size distribution on the screen has an average diameter of 54.0 (SD 52.3) micrometers, and a minimum detectable size of 7 micrometers (to determine these parameters, stains are treated like disks of the same area). These values are very close to the ones reported in an analogous experiment performed to characterize actual coughing by Duguid [45], where the average stain size was 53.4 micrometers. Even when the stains have larger radii than the droplets causing them, it is safe to say that we are basically detecting the stains associated to ballistic droplets at the exit of the nozzle, i.e., those bigger enough to move as projectiles between the source and the receptor.

It is worth noting that the electronic parts involved in the setup can be purchased for 50 USD or less, while the (detergent) bucket, spray bottle and scanner are not difficult to find in homes or offices. Finally, the image processing code was written by the authors –and can be easily coded on several free platforms. This illustrates the “frugal” character of our proposal. Sec. S2 of supplementary material describes the experimental apparatus and procedure in more detail. Furthermore, our setup allows measuring the droplets that actually hit the “face” of the receiver, instead of visualizing a “cloud” of droplets in the air [11]. This might be seen as a practical advantage.

## RESULTS AND DISCUSSION

### Relative quantification of the ballistic protection of different cloths

The main goal of our proposal is to compare the ability to block ballistic droplets by different cloths available to fabricate face masks, in order to select the most effective one based on a quantitative criterion. So, we are not proposing an absolute, but a relative evaluation protocol.

In Fig. 2 we show micrographs and stains patterns corresponding to the blocking materials under study: a neck gaiter (labeled Gaiter), artificial silk (Silk), a cotton handkerchief (Handkerchief), cotton tablecloth (Table Cloth) and surgical gown (Gown). In addition, we also include the stain pattern without any blocking material. It becomes clear that all materials deployed in CONF1 are able to block most of the ballistic droplets emitted by the nozzle.

**Figure 2.**
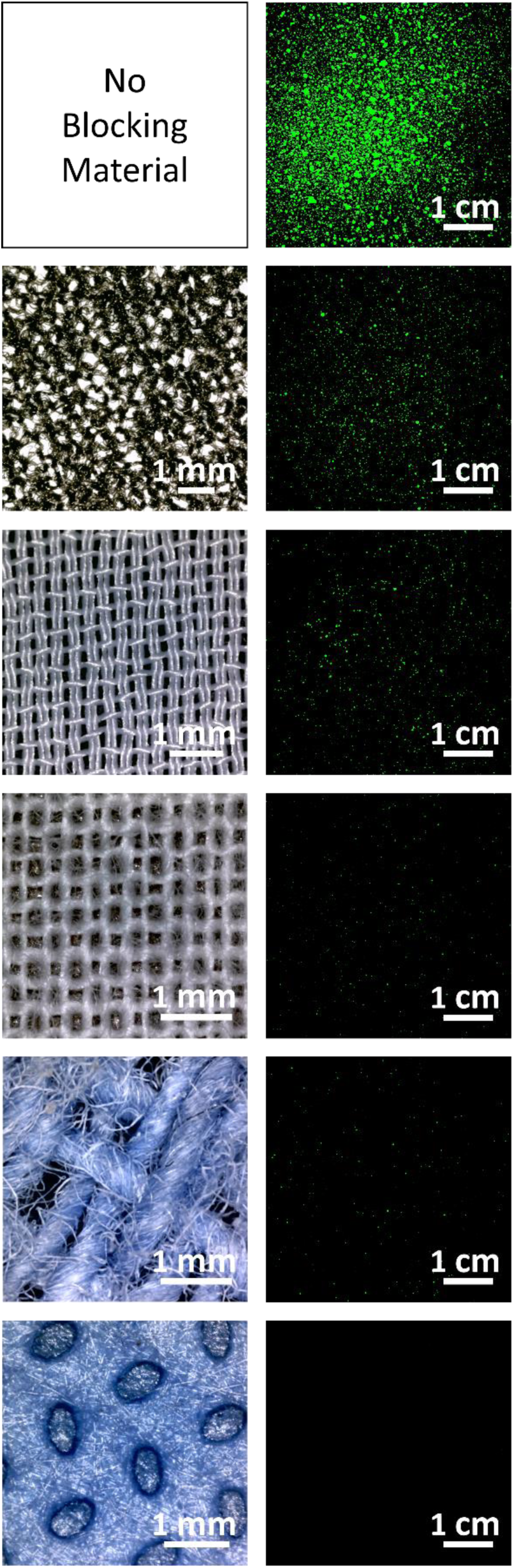
Micrographs of blocking materials and their stains patterns. From top to bottom: No blocking material, Gaiter, Silk, Handkerchief, Table Cloth and Gown. For no blocking material the pattern corresponds to CONFFREE and for the rest, patterns correspond to CONF1. Stains have been artificially colored to aid visibility.

In order to quantify the capacity of a given mask material to isolate a receptor from the ballistic contamination emitted from a source, we propose a parameter named Ballistic Blocking Capacity, defined as:

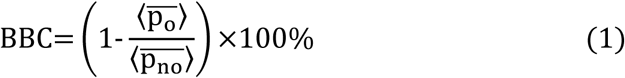

where 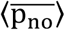 (standing for *pixels no obstacle*) is the average of the pixel values in the image corresponding to the screen without obstacle, averaged over the images from all similar experiments, and 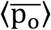 (*pixels obstacle*) is the analogous magnitude when an obstacle is deployed. Since both averages are within the interval [0,1], BBC = 100% when the obstacle has stopped all droplets 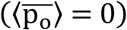, and BBC = 0% when all droplets have managed to pass through the obstacle 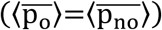.

The upper panel of Fig. 3 reports the 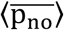 value and the 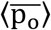 values for every material, which are inserted in Formula (1) in order to calculate the corresponding BBC values for a given configuration. The lower panel shows those values for materials under study deployed in CONF1. Three repetitions were made for each type of experiment.

**Figure 3.**
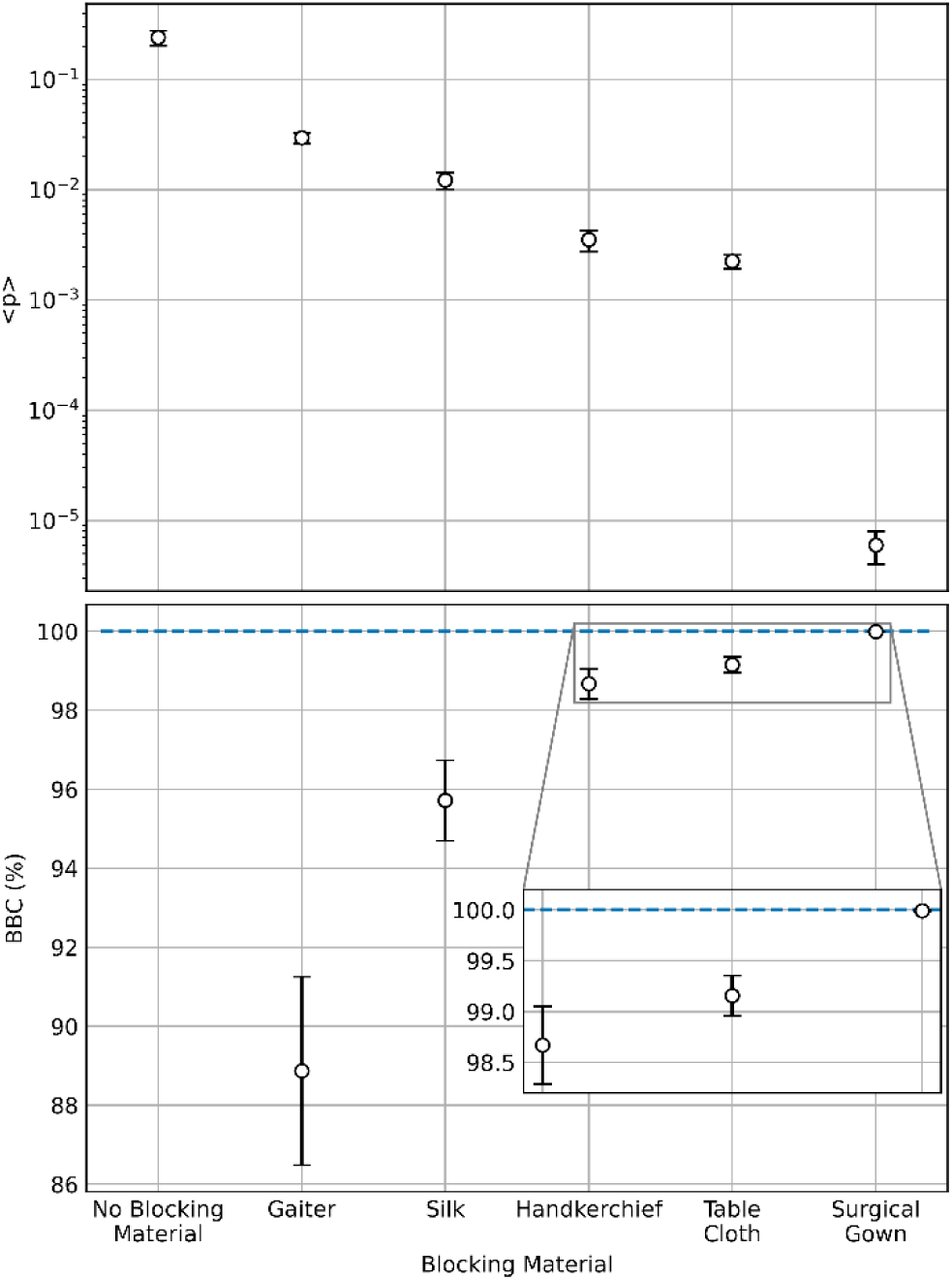
Ballistic blocking for various fabrics. (a) Values of 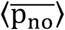 for no blocking material in CONFFREE, and of 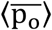 for different blocking materials in CONF1 (notice that both 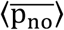 and 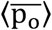 have been labeled as ⟨*p*⟩). (b) BBC for the same materials, calculated through formula (1). Inset: zoom near BBC = 100%. See supplementary material Sec. S3 for uncertainty analysis.

The decrease of the BBC values from Gaiter to Gown may be seen as a way to validate our protocol, since the results are partially analogue to previous studies using other methods to assess the protection ability of materials used in homemade face masks (see, for example, [11,17]). However, we do not claim that BBC is an absolute measure of the blocking capacity of a given material: our protocol aims at the quantitative comparison of the blocking ability of different materials.

### Further experimental geometries

#### a) Configurations mimicking real life

It is possible to establish a link between different experimental configurations and situations observed in real life, where a sneezing, coughing or talking individual (source) expels mucosalivary particles, eventually hitting the face of another person (target). As sketched in Fig. 4(a), CONF1 mimics a situation where the face of a masked target is hit by the ballistic emissions of a maskless source located 19.0 cm away from her/him –the choice of distance is within the face-to-face proximity range measured by Zhang *et al*. [42], which can be easily experienced during rush hours in a bus in Havana, a Metro in Paris or a subway in New York. Fig. 4(b), labeled CONF4, illustrates the inverse situation: the target is maskless, while the source is wearing a mask. Finally, Fig. 4(c) depicts a configuration called CONF1+4 where both the source and the target are wearing masks. The three situations can be arguably found in real life scenarios.

**Figure 4.**
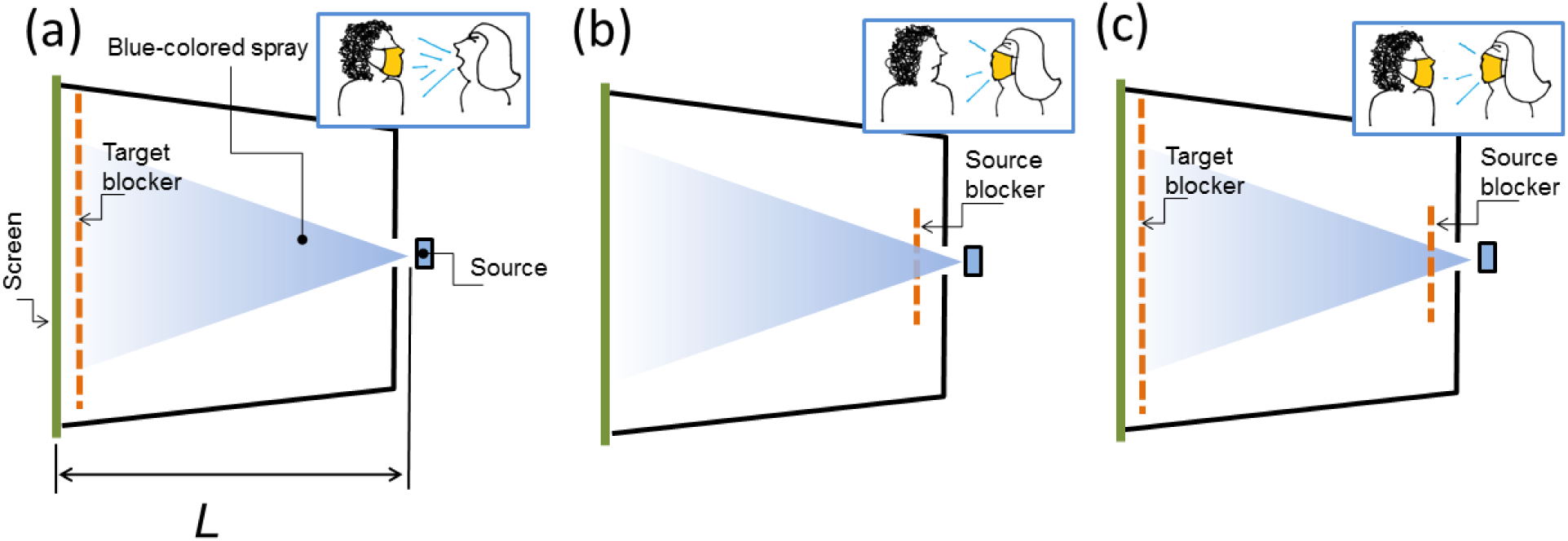
Configurations mimicking real life. (a) CONF1: The target protects herself (himself) using a mask, while the source is unmasked. (b) CONF4: Only the source wears a mask. (c) CONF1+4: Both the source and the target are wearing masks.

For evaluating the effectiveness of a mask near the face of the receiver against ballistic droplets produced by the emitter in each of the three scenarios described above, we insert in (1) the calculated values of 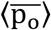 corresponding to experiments performed in configurations CONF1, CONF4 or CONF1+4. Fig. 5 shows the results: the BBC values for CONF1 are systematically smaller than those corresponding to CONF4. To test if the observed difference is statistically significant for a given blocking material, a one-tailed Mann-Whitney U-test was performed. In all cases the null hypothesis was rejected within a 95% confidence level, proving that BBC values for CONF4 are statistically larger than those for CONF1. In other words: the perception that a mask wearer protects other better than her/himself is true for the case of ballistic droplets.

**Fig. 5.**
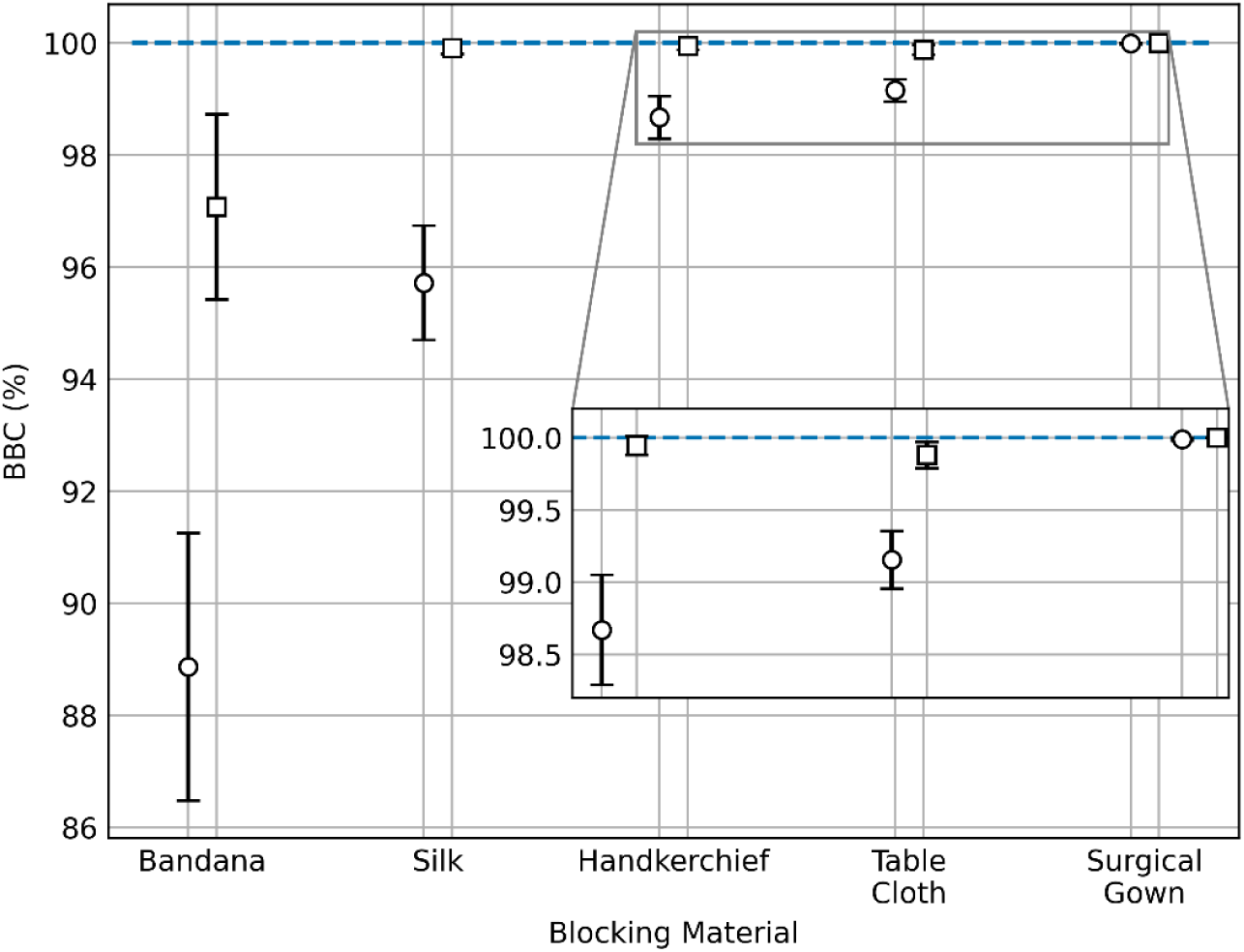
A target is better protected by a mask wore by the source than by a mask wore by her/himself. BBC for different mask materials in CONF1 (circles) vs CONF4 (squares). Inset: zoom near BBC = 100%.

Finally, we measured the BBC value in CONF1+4 for Silk, giving a value of 99.99 ± 0.01 %. This confirms the intuitive notion that the best protection is achieved when both the source and the target are wearing masks. However, the protection is not perfect, at least for the case of relatively small face-to-face distances examined here.

### b) Intermediate configurations

In order to further explore the phenomenology of how ballistic droplets are blocked by cloths, we performed experiments where a single sample of Silk was deployed at two extra positions between the source and the target. They are sketched in Fig. 6(a). Fig. 6(b) shows the dependence of the calculated BBC values as a function of the distance between the blocking material and the nozzle. As expected from the previous measurements for CONF1 and CONF4, BBC decreases with the distance from the nozzle. What is somewhat unexpected is that the decrease is well fitted to a linear dependence, as represented by the blue line in Fig. 6(b).

**Fig. 6.**
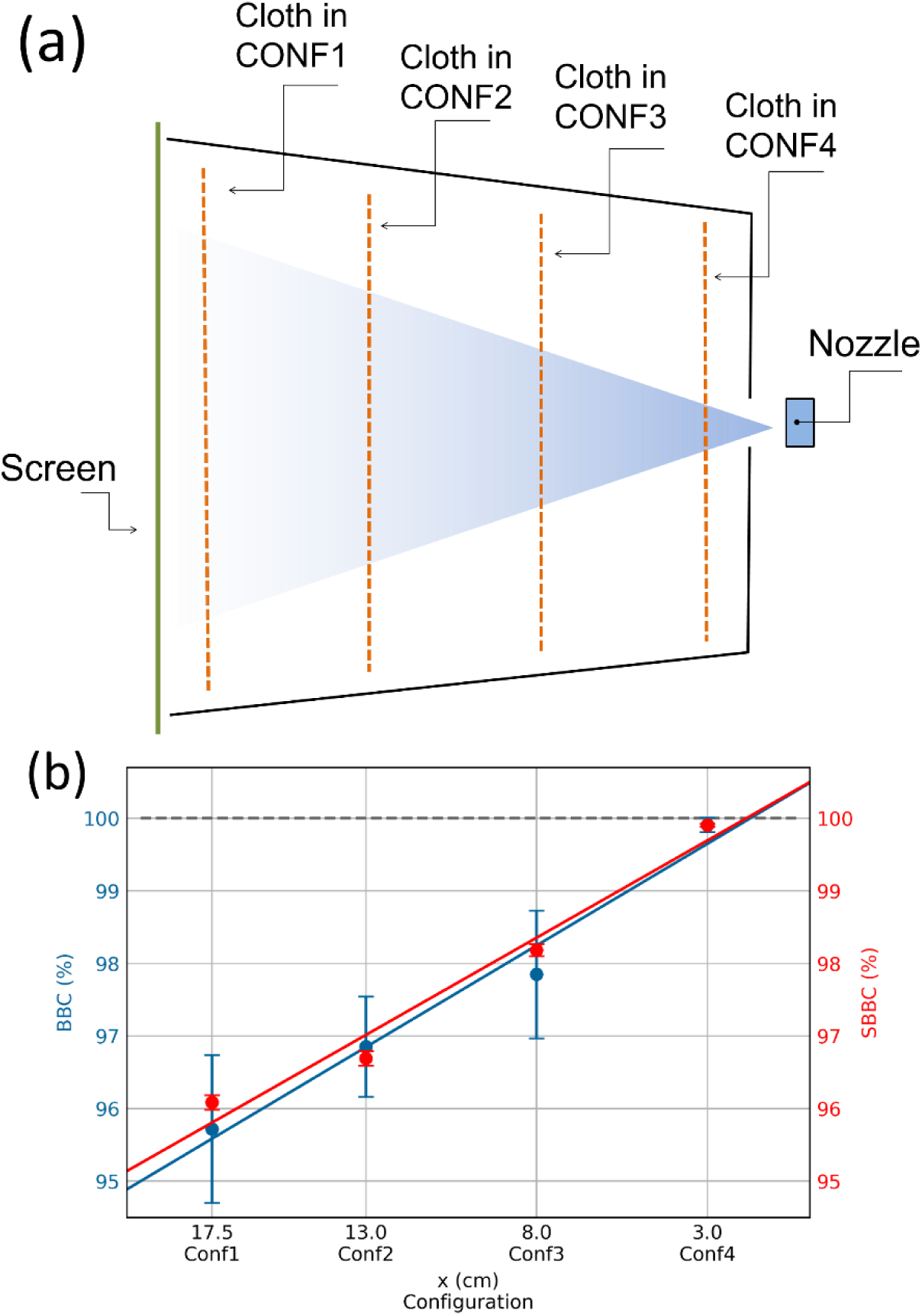
BBC and SBBC for intermediate positions of the blocking material. (a) Sketch of the cloth positions under study. (b) Experimental (BBC) vs. model (SBBC) results. Blue and red points correspond to experimental data and model calculations, respectively. Blue and red lines correspond to linear fits to the experimental and theoretical points, respectively.

In an attempt to understand this behavior, we propose a simple mechanical model, as follows. Droplets are assumed to move ballistically –i.e., like projectiles– from the source to the receptor. Along the trajectory, there are two basic scenarios. (a) As the droplets move through the air, they just feel the gravitational force and the viscous drag due to the interaction with the air and (b) During the interaction with a blocking cloth, droplets are able to pass through it with a certain probability; in the case they emerge from the opposite side, their velocities would decrease. The step-by-step implementation of the model in order to mimic the actual experiments is described below.

Let us assume a droplet of radius r emerging from the nozzle with velocity of magnitude v_0_ that makes an angle θ_0_ with the horizontal direction. If the nozzle is located at (x_0_, y_0_) and the only forces acting on the droplet after ejection are the gravity 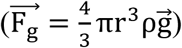 and the Stokes drag 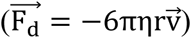 associated to the interaction with the air [46], it follows a trajectory given by

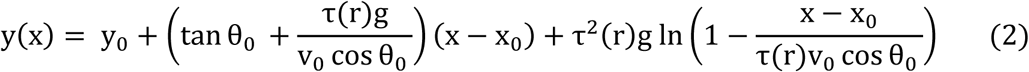

where g = 9.81m/s^2^. Relaxation time τ(r) = 2ρr^2^/9η is the time it approximately takes a droplet of radius *r* to reach its terminal velocity v_T_ = τ(r)g = 2ρr^2^g/9η. We assumed η = 18.12 × 10^−6^Pa · s (absolute viscosity of the air at 20^0^ C) and ρ = 995.7 kg/m^3^ (density of water at 30^0^C) [46].

The interaction of a flying droplet with a given blocking barrier is modeled by means of two dimensionless parameters. Firstly, the *tunneling probability* P ∈ [0,1], which is the chance to pass through the obstacle (so 1 − P is the probability to be blocked). Secondly, the *deceleration factor*, f ∈ [0,1] defined as the ratio between the droplet velocity moduli as it exits and enters the blocking element, respectively. Notice that in our model, neither P nor f depend on the droplet radius, or on its angle of incidence as it penetrates the obstacle.

Each simulation involved 100 000 droplets emitted by the source, whose radii were generated from a size distribution extracted from an experiment where the screen was located 4 cm away from the nozzle, with no blocker in between (see Experimental section). All generated droplets were assumed to start moving with a velocity of magnitude v_0_ = 3 m/s, extracted from the experiment, but each droplet is emitted following a random direction within an angular range [−10°, 10°] around the horizontal (see supplementary material Sec. S1).

In order to compare with the experimental BBC values, we define the Simulated Ballistic Blocking Capacity (SBBC), which is also defined by formula (1), where 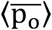 and 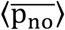 are computed as the total droplet volumes reaching the center of the screen when an obstacle is deployed or not, respectively.

We used P and f as free parameters to generate SBBC values as close as possible to BBC values for each of the four positions of the cloth by minimizing the weighted sum of squares 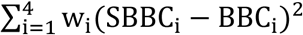 w_i_(SBBC_i_ − BBC_i_)^2^, where i = 1, 2, 3 and 4 represents the four positions of the cloth and *w*_*i*_ = 1/*σ*_*i*_^2^; *σ*_*i*_^2^ being the variance of BBC for the i-th configuration. The red points in Fig. 6(b) shows the results, corresponding to p = 0.04 and f = 0.56. The red line in the same graph is a linear fit to the theoretical values. It is worth noting that the linear extrapolations corresponding to BBC and SBBC values larger than 100% should not be taken into account.

So, both the experimental results and the output of our model show linear dependences of the blocking capacity vs. cloth position within our range of p and f, and the difference between both slopes is as small as a 4.3 %. It means that a minimalistic model where droplets move ballistically submitted only to gravity and air drag, and interact with the cloth in a simple way is able to reproduce the experimental results accurately.

## CONCLUSIONS

By using frugal apparatus and a relatively simple experimental protocol, we have been able to evaluate the relative blocking capacity against ballistic droplets of some materials commonly used in the fabrication of face masks at home. This method could be useful in situations where it is necessary to compare the performance that available materials would have as a face mask, but aerosol-sensitive techniques are not available [9, 15, 16, 20, 22, 23]. It is worth mentioning that one of these papers also study the blocking capacity of face masks at short distances (< 50 cm) [16]. In spite of not dealing with jet-like emissions, they arrive to results consistent with ours.

Since our simple apparatus easily allows testing cloths in different positions, we are able to evaluate their performance in “source control” and “wearer protection” modes. Concentrating on the study of ballistic droplets, our results show that a cloth mask worn by an infected subject is effective to avoid spreading the disease, while a healthy person is less protected from an external infection source when wearing the mask, if others are not wearing it. This is consistent with very recent experimental results using more complex apparatus able to detect aerosols [21-23]. We also show that a larger protection is achieved when both the source and the receiver wear masks, although it does not reach a 100%, even for relatively large, ballistic droplets.

Finally, we are able to reproduce the blocking capacity of a given cloth located at different positions between source and target by using a simple mechanical model where the droplets behave as projectiles experiencing gravity and air drag, and the effect of the cloth is characterized by just two phenomenological parameters.

## Data Availability

All data produced in the present study are available upon reasonable request to the authors

## Supplementary material

See the supplementary material for visualization of the free jet and determination of its front’s velocity, image acquisition and process details, and uncertainty analysis.

## Acknowledgments

P. G. Gil is acknowledged for suggesting home-made masks as an urgent and important subject of study. T. Shinbrot contributed with key suggestions and revision of the manuscript. N. Martínez coordinated lab access a number of times during home self-isolation period. P. Altshuler-Rivera is dearly thanked for collaborating in spray visualization experiments at home.

## Data Availability

The data that support the findings of this study are available from the corresponding author upon reasonable request.

