## Supplementary material for "Relative assessment of cloth mask protection against ballistic droplets: a frugal approach"

### S1. Visualization of the free jet

The jet was visualized by illuminating with a light "cone" produced by a green laser (λ = 532 nm and peak power of 50 mW). The images were acquired at 30 fps using a NIKON D5100 camera. Figure S1 illustrates the evolution of the jet based in its visualization.

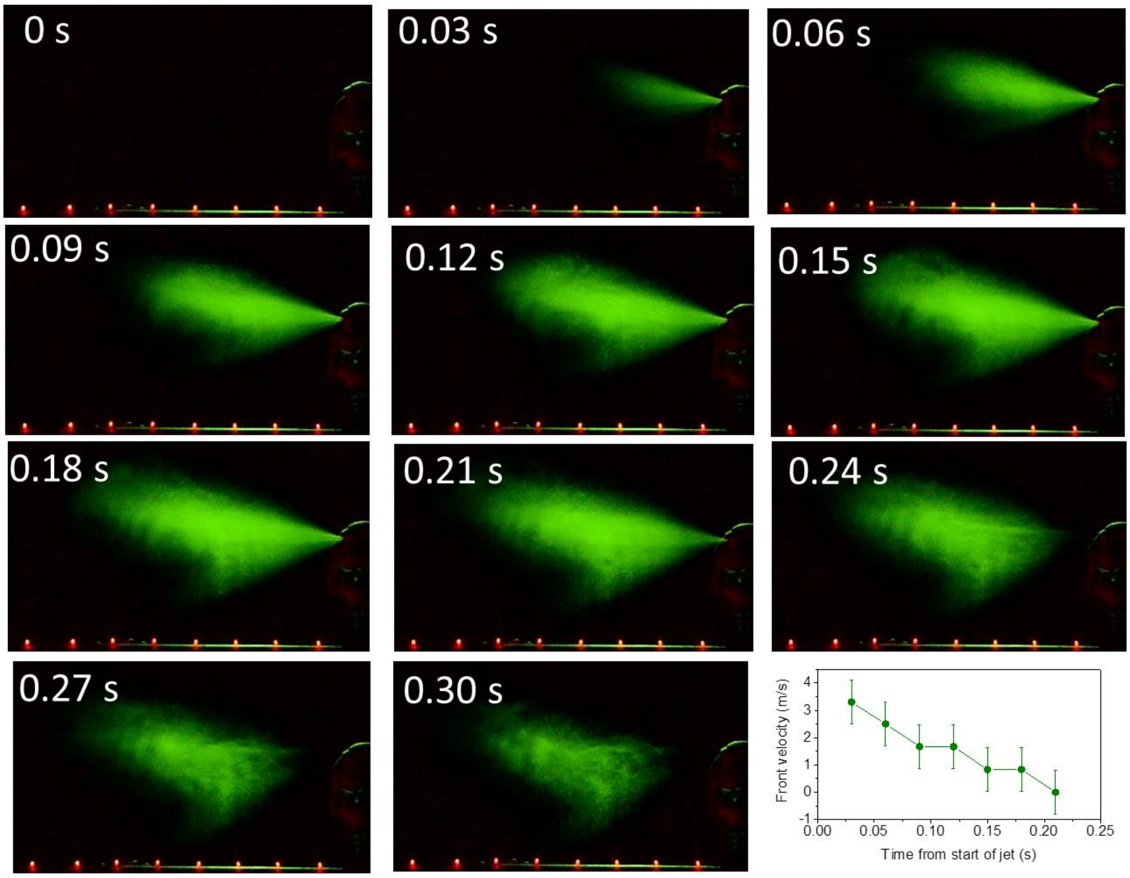


**Fig. S1 Visualization of the free jet.** The 11 photographs are a time sequence of snapshots of the free jet, visualized using laser light. The time count starts when the nozzle starts generating liquid. In each of the snapshots, the green light indicates the presence of liquid particles emitted by the nozzle (located at far right within each image). The red spots are LEDs located at a distance of 5 cm from each other. Bottom right panel: velocity of the jet front as time goes by. (See Multimedia view, where the video has been slowed down approximately 7 times for better visualization).

### S2. Experimental details

#### Image acquisition and processing

75 g/m$^2$-density paper sheets containing the stain patterns were scanned with a resolution of 3200 dpi and 24-bit RGB color, using a scanner EPSON model Perfection V370 Photo. Then, the image was negated, and its red component binarized. As the spots are blue,

this component provided the best contrast between the stains and the background. After careful comparison between the original patterns and the binarized images, we chose a binarization threshold of 0.3.

If the image with the stain pattern was subtracted from the scan of that very same sheet of paper made before the droplets were shot (in order to account for intrinsic lack of homogeneity of the paper), the results for BBC were negligibly different.

**Micrographs of blocking materials**

Micrographs of different blocking materials were obtained using a Dino-lite Premier digital microscope.

### S3. Uncertainty analysis

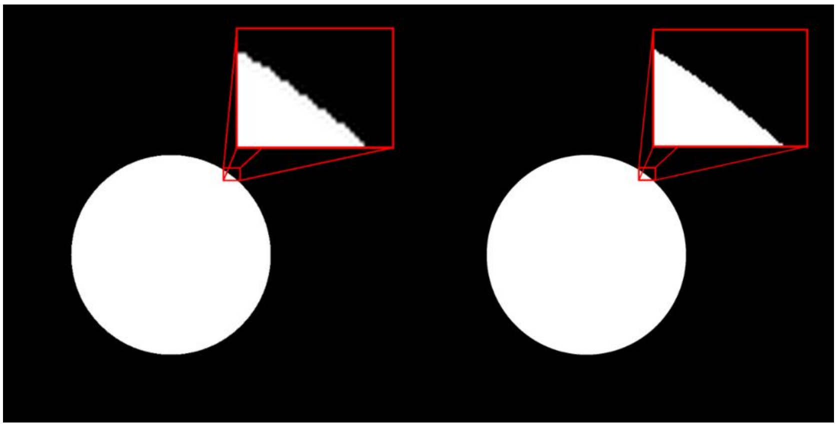

Fig. S4 Uncertainties in image digitizing. Two raster images created from the same white disk on a black background vector graphics, simulating the stains in the binarized images of the experiments. Left panel: 3200 dpi resolution image. Right panel: 6400 dpi resolution image. In the insets it can be seen the difference between the analog regions in the disks boundaries.

In the calculation of $\langle\overline{p_{no}}\rangle$ and $\langle\overline{p_o}\rangle$ (both referred as $\langle\overline{p}\rangle$ from now on, as they are computed in the same way), there are two sources of Type B standard uncertainty: the stained paper scan resolution and the finite precision with which the program reads the pixel values.

In the case of the former, a test was made to check if an increase in the image resolution would affect significantly the values of $\langle\overline{p}\rangle$ calculated in the experiments. In order to do so, two raster images were created from the same vector using resolutions of 3200 and 6400 *dpi* and $\langle\overline{p}\rangle$ was calculated on both of them. The difference between both was around 0.1% of the value of the 3200 *dpi* image, so the influence of this source of uncertainty can be neglected as compared to the influence of the fluctuations between repetitions of the same experiment, as we will see.

The program used for image processing reads by default the pixel values with a precision of 15 significant decimal digits. As the pixel values are within the interval [0,1] the difference between two pixels whose values differ up to R = $10^{-15}$ can be detected. Due to binarization, this resolution would only affect the pixels whose values are within the interval $[\mathrm{Th}-\mathrm{R}, \mathrm{Th}+\mathrm{R}]$ being $\mathrm{Th}$ the binarization threshold used in the experiments. In a typical image, the proportion of pixels that satisfies this condition varies from zero to $10^{-15}$, so the influence of that source of uncertainty can be also neglected. In summary, the influence of Type B is negligible compared to the Type A standard uncertainty on the evaluation of $\langle\bar{p}\rangle$'s combined standard uncertainty.

For all experimental configurations, $\langle\bar{p}\rangle$'s Type A standard uncertainty was computed using the well-known positive square-root of the unbiased estimator for the variance of a sample divided by its length. Each experiment was repeated 3 times and a 95% confidence interval was selected for the calculations, so according to Eq. (1), the formula used for the expanded uncertainty of BBC is

$$\mathrm{u(BBC)} = \mathrm{t}_2\left(\frac{1}{\langle\overline{\mathrm{p}_{\mathrm{no}}}\rangle^2}\mathrm{u_A}^2(\langle\overline{\mathrm{p}_{\mathrm{o}}}\rangle)+\frac{\langle\overline{\mathrm{p}_{\mathrm{o}}}\rangle^2}{\langle\overline{\mathrm{p}_{\mathrm{no}}}\rangle^4}\mathrm{u_A}^2(\langle\overline{\mathrm{p}_{\mathrm{no}}}\rangle)\right)^{1/2} \tag{S1}$$

where $\mathrm{t}_2 = 4.30$ is the factor corresponding to the confidence interval of choice and the effective degree of freedom (3 experiments were made for every fabric and configuration).
